# A systematic review of the impact of the Alpha and Gamma variants of concern on hospitalization and symptomatic rates of SARS-CoV-2

**DOI:** 10.1101/2021.08.13.21261151

**Authors:** Alexandra Schroeder, Matthew R. MacLeod

## Abstract

The hospitalization and symptomatic rates of Severe Acute Respiratory Syndrome Coronavirus Disease 2019 (SARS-CoV-2) are key epidemiological parameters affecting risk analyses conducted for the Canadian Armed Forces (CAF) during the Coronavirus Disease-19 (COVID-19) pandemic. As one of the criteria of a variant of concern (VOC) is that it affects disease severity, the authors sought to understand whether the Alpha and Gamma VOCs are significantly different in these two parameters than the original wildtype of SARS-CoV-2, the most prevalent in relevant areas of Canada as of study initiation. Searches for studies were conducted in Scopus and PubMed, and located through following citations and receiving studies from daily literature scans. For the hospitalization outcome, effect ratios relative to original wildtype were included. For the symptomatic ratio, the ratio itself for each variant was used. Analysis of age-related effects was of particular value, as CAF members are primarily adults under the age of 60. The firmest conclusion of this review is that the Alpha VOC comes with a higher relative risk of hospitalization compared to the original wildtype, most likely above 1.4, while unlikely to be above 2, with the balance of evidence being that the relative risk is not significantly modified by age. The evidence for Gamma is more limited, but the odds ratio may be above 2, and potentially much greater than that, especially for those 20-39 years of age. For both VOCs reports on symptomatic ratio differed on whether there was an effect, as well as its potential direction.

## 1. Introduction

When this review was launched, relatively few studies or reviews were available on the impact of Alpha and Gamma variants of concern (VOCs) of Severe Acute Respiratory Syndrome Coronavirus Disease 2019 (SARS-CoV-2) (1) on hospitalization and symptomatic rate. A February 2021 research note on the severity of Alpha was available (2), citing primarily unpublished sources from the United Kingdom, and largely focusing on mortality. A research brief was available online combining elements of a meta-analysis with original results for Ontario, Canada, of the combined effect of all VOCs circulating as of March 2021 on hospitalization and intensive care unit (ICU) admission (3). The authors could not locate a review of symptomatic ratio for either VOC, nor one of hospitalization for Gamma. Estimates of the increase in transmission rate for VOCs are more easily inferred from publicly available data (e.g., (4)), whereas reliable data on these two parameters of interest is more difficult to collect; the challenges of creating such estimates for disease severity were noted in a *Lancet* commentary (5). As such, we initiated a systemic review following the Preferred Reporting Items of Systematic Reviews and Meta-Analyses (PRISMA) checklist (6).

We conducted the review with the intent to inform the Canadian Armed Forces (CAF), which guides its design. Hospitalization was selected as the severe outcome of most concern as CAF members are at low risk of mortality from SARS-CoV-2 infection due to their age and low level of comorbidities (7), but some members have been hospitalized by infection – which for instance creates operational concerns with respect to a potential requirement for medical evacuation (8). Symptomatic ratio is of concern as it drives both the likelihood of an individual being ‘missed’ as an infection when departing for an operation or exercise (9), and the probability that individuals are identified for self-isolation once in an environment where infection is spreading (see e.g., (8)). The study participants of most interest are adults aged 18-60, as the standard retirement age in the CAF is 60. The exposure of interest is to Alpha or Gamma VOC as compared respect to original wildtype SARS-CoV-2 virus, as these were the most predominant VOCs in communities of interest to the CAF at the time the study was initiated. The outcomes of interest are the likelihood of hospitalization and likelihood of developing symptoms, including any mediation by age. Cohort studies and cross-sectional studies are included.

## 2. Methods

A formal review protocol was not created and registered. The authors followed the PRISMA checklist (6), making use of the Covidence systematic review manager (10).

The primary outcomes of interest were likelihood of becoming symptomatic and/or being hospitalized given infection with SARS-CoV-2, looking for differences between the original wildtype and the Alpha and/or Gamma VOCs. Of particular interest were studies that presented effects by age, given the focus on adults below the age of 60, and previously observed age-related effects of the virus (7). The currently accepted World Health Organization labels were not yet announced when the study was initiated, so Alpha was primarily known by its Pango lineage B.1.1.7, and Gamma as P.1 (1). Given that Alpha was not recognized and designated as a VOC until December 2020, and Gamma until January 2021 (1), studies conducted entirely 2020 were assumed not to contain sufficient data of interest. Studies in English and French were considered. Both pre-prints and published papers were included, given the important role pre-prints have played in understanding the pandemic (11).

The primary search was conducted on Scopus (12) and PubMed (13) on 21 May 2021. The authors became aware of additional studies through their contacts with the Public Health Agency Canada (PHAC) external modelling group, a daily scan of Coronavirus Disease-19 (COVID-19) publications distributed by PHAC, an Ontario Science Table report (3) which referred to pre-prints not in the initial search, as well as papers that were referenced in papers subjected to full-text review.

The following string was used for the database searches:

~~~
(“COVID”) OR (“COVID-19”) OR (“SARS-CoV-2”) AND (“B.1.1.7” OR
“VOC 202012/01” OR “501Y.V1” OR “P.1” OR “B.1.1.28”) AND
(“SEVERE” OR “SEVERITY” OR “HOSPITALIZATION” OR
“HOSPITALISATION” OR “HOSPITAL” OR “DEATH” OR “ASYMPTOMATIC”)
~~~

We screened all retrieved titles and abstracts for eligibility and then full texts were screened if relevant. Disagreements on the eligibility of articles were resolved through discussion. Articles were excluded if they were:

1. A preprint of another article retrieved in the search,
2. A Review Article, Commentary Article or Case Study,
3. Had no information on the Alpha or Gamma variants of concern,
4. Of the wrong age group (not 18-60 or some close approximation),
5. Contained the wrong outcome (no information on the risk of hospitalization or symptomatic ratio),
6. Were of participants with co-infection of VOC with another SARS-CoV-2 lineage,
7. Had no isolated analysis of the effect of VOC B.1.1.7 or P.1,
8. Had a tentative outcome ratio, but data too time limited to capture severe outcomes; or
9. Had descriptive statistics only, with no effect estimate.

Data from each article was extracted independently by each author. Data extraction conflicts were resolved through discussion between the two authors. The following information was extracted from each article: title, study identification, date published, authors, country in which the study was conducted, aim of study, highlighted SARS-CoV-2 variants, study design, start date, end date, study funding sources, possible conflicts of interest for study authors, total number of participants, population description, age group, age-related results, gender-related results, risk of hospitalization/critical care admission, symptomatic ratio, and other information on the severity of the Alpha or Gamma VOC.

To assess the risk of bias of the included studies, we used the cohort studies version of the Newcastle-Ottawa Scale (NOS) for assessing the quality of nonrandomised studies in meta-analyses (14). This was done at the outcome level, as studies tended to use lower quality methods when reporting on symptomatic ratio.

The preferred summary measure of interest for hospitalization was the risk ratio of the variant (Alpha or Gamma) compared to that of the original wildtype; where this was not available, odds ratio or hazard ratio was used. The preferred summary measure for the symptomatic ratio was the ratio itself, whether for the variant or the original wildtype.

The authors did not perform a quantitative meta-analysis. For Gamma there was insufficient data. For Alpha, the authors determined a qualitative summary would be more appropriate, as three different measures of relative effect were used across only six studies reporting on hospitalization, and the reporting on symptomatic ratio was even less consistent. There is also a risk of over precision when combining preliminary studies – e.g., an earlier meta-analysis of two studies, of Alpha’s relative risk of hospitalization (3) found near perfect agreement, but later versions of those studies reported estimates that had moved in opposite directions.

The primary risk of bias across studies is that the VOCs had very different patterns of geographical circulation; the result is that studies are biased towards both where they circulated, and also the capacity and incentives of researchers within the public health systems in those areas to conduct studies and publish them. In particular, Alpha heavily affected the United Kingdom (along with several countries in Europe), where many of the studies cited here originated, which implies the conclusions may be impacted by the characteristics of its population and of its healthcare system. Gamma is comparatively less studied in this respect. Outcomes could be expected to improve over time as treatment protocols evolved, and perhaps as some of the most susceptible and vulnerable individuals were infected early in the epidemic; that said, several studies were conducted while multiple variants were circulating contemporaneously, mitigating this risk. Given the crisis induced by the pandemic, there may also be publication bias towards reporting results perceived as presenting significant risks to clinical or public health, whereas findings of little to no effect may be delayed for later publication, if they are published at all.

No additional analyses were pre-specified. However, where the data was available in an included study to calculate an outcome of interest which the original authors did not, this was pursued. In particular, one included study presented the percentage of total hospitalizations at a given time within each age group, rather than the percentage of individuals within each age group who were hospitalized.

## 3. Results

237 studies were screened, of which 4 were ultimately included, in addition to 5 additional studies that were identified through additional follow-up, for a total of 9. The full PRISMA flow diagram is included in Figure 1.

**Figure 1.**
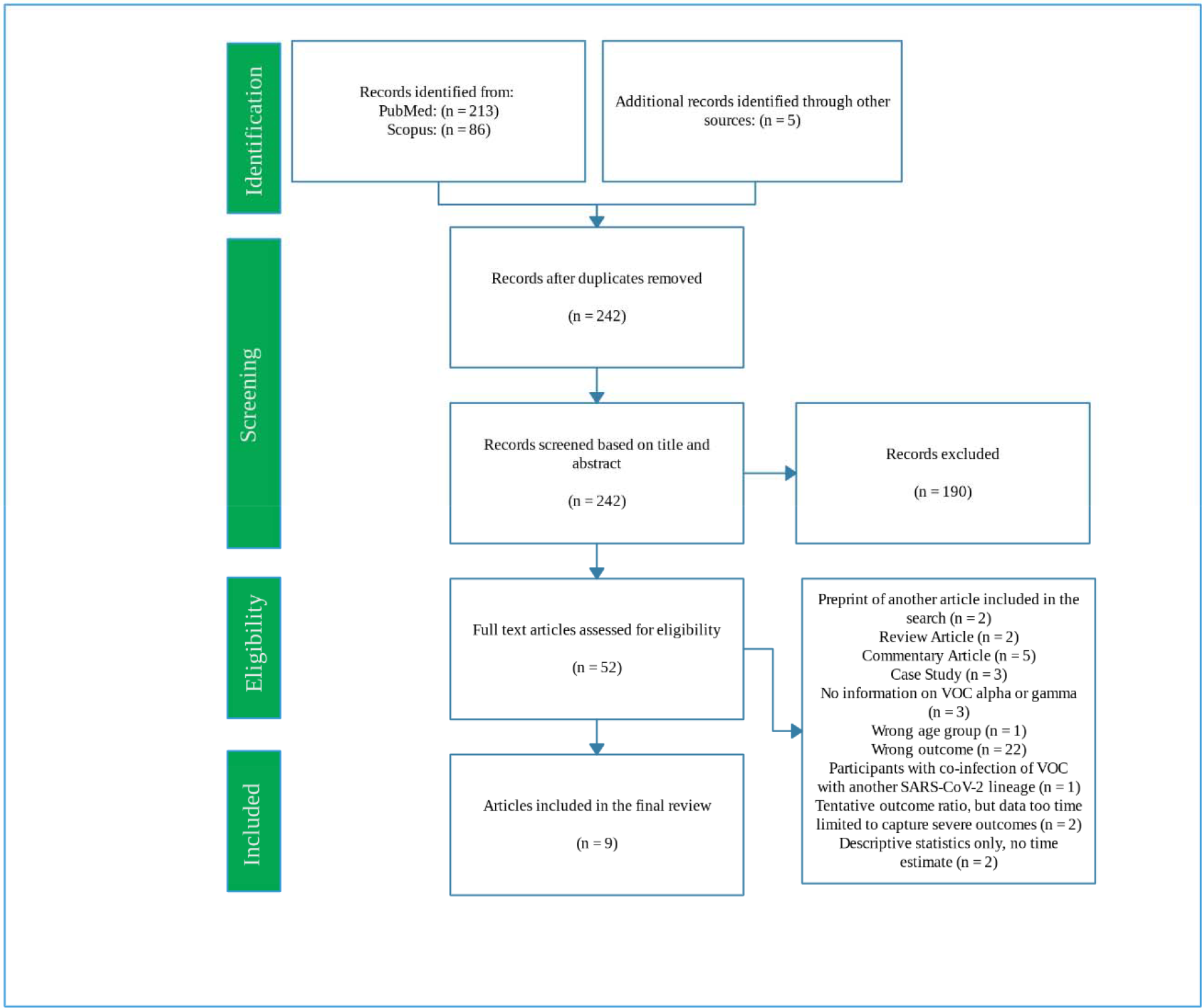
Preferred Reporting Items of Systematic Reviews and Meta-Analyses flow diagram.

Of the included studies, four were conducted in England (15-18), and will be numbered in the tables that follow. One study was conducted in Denmark (19), one in Ontario, Canada (20), one in Southern Italy (21), and one in Northwest Spain (22). One includes data from seven European national surveillance systems (23). The exposure and comparison are between original wildtype SARS-CoV-2 and the relevant VOC, or some suitable proxy (e.g., S-Gene Target Failure (24)), with the outcomes of interest being hospitalization and/or the development of symptoms. All studies were cohort studies. Results for Alpha and Gamma are reported separately below.

### 3.1 Alpha

The outcome presented in the most studies (n = 6) was the relative hospitalization likelihood for Alpha compared to wildtype, using three different outcome measures – risk ratio (RR), odds ratio (OR), and hazard ratio (HR), all of which showed a statistically significant increase in the likelihood of hospitalization with Alpha; these are summarized in Table 1. The one study presenting the preferred summary statistic of risk ratio reported 1.42 (95% Confidence Interval: 1.25, 1.60), and did not observe a statistically detectable differential impact by age (19). Two studies from England reported an adjusted hazard ratio of 1.99 (1.5, 2.4) (15) and 1.52 (1.47, 1.57) (16). While (15) found no significant interaction with age group (p = 0.15), (16) did, reporting for CAF age groups of interest:

**Table 1:** Summary of hospitalization outcomes for Alpha VOC compared to wildtype.

| Location | Participants | Relative Effects<br>(95% CI) | Risk of bias (Newcastle-Ottawa Scale) |  |  |
| --- | --- | --- | --- | --- | --- |
|  |  |  | Selection | Comparability | Exposure |
| Denmark (19) | 50 958 | **RR 1.42** (1.25, 1.60) | ◇◇◇◇ | ◇◇ | ◇◇◇ |
| Ontario (20) | 216 678 | **OR 1.74** (1.62, 1.86) | ◇◇◇◇ | ◇◇ | ◇◇◇ |
| Europe (23) | 23 343 | **OR 1.6** (1.2, 2.3) | ◇◇◇◇ | ◇◇ | ◇◇◇ |
| Southern Italy (21) | 3075 | **OR 3.44** (1.76, 6.75) | ◇◇◇◇ | ◇◇ | ◇◇◇ |
| England 1 (15) | 201 852 | **HR 1.99** (1.5, 2.49) | ◇◇◇◇ | ◇◇ | ◇◇◇ |
| England 2 (16) | 839 278 | **HR 1.52** (1.47, 1.57) | ◇◇◇◇ | ◇◇ | ◇◇◇ |

- 20-29: 1.30 (1.19, 1.42)
- 30-39: 1.41 (1.32, 1.51)
- 40-49: 1.59 (1.50, 1.69)
- 50-59: 1.58 (1.50, 1.67)

Three studies reported odds ratio, which well approximates the risk ratio for rare diseases, but will overestimate positive associations. The study in Ontario reported on N501Y+ variants relative to non-VOC (20), which includes both Gamma and Beta cases as well as Alpha (1), reporting 1.74 (1.62, 1.86) as the odds ratio between N501Y+ VOC and original wildtype. As Alpha made up more than 90% of N501Y+ variant cases in Ontario at all times during the study period (25), this estimate can be assumed to be dominated by Alpha. A report using European surveillance data from Cyprus, Estonia, Finland, Ireland, Italy, Luxembourg and Portugal found an overall adjusted odds ratio of 1.6 (1.2, 2.3) or 1.7 (1.0, 2.9), for Alpha, depending on the choice of technique (23). They report a statistically significant modification of adjusted odds ratio by age, giving for CAF age groups of interest:

- 20-39: 3.0 (1.4, 6.8)
- 40-59: 2.3 (1.0, 5.4)

Finally, a study using data from Southern Italy estimated an adjusted odds ratio of 3.44 (1.76, 6.75) for hospitalization for Alpha with respect to original wildtype (21), and did not directly report on the presence or absence of an age trend.

Five studies reported on the symptomatic ratio for Alpha, as reported in Table 2, three of which were also included in Table 1. The first additional study is a United Kingdom (UK) Office of National Statistics report issued amidst the major Alpha outbreak in January 2021 (18), finding that those infected with Alpha were statistically significantly more likely to self-report being symptomatic at 52.92% (51.27, 54.56), compared to those with the original wildtype at 41.67% (40.05, 43.30); of specific symptoms, statistically significant differences were observed with those with Alpha being more likely to report cough, while less likely to report loss of taste or loss of smell (18). An ecological study in England using self-reports from an application (17) found no significant effect across 14 specific symptoms. The aforementioned European surveillance report found a significant difference between symptomatic rate for Alpha of 72.6% compared to 81.4% for original wildtype (23). The study of Southern Italy noted above found a statistically significant difference in symptomatic infection, reporting 51.7% of Alpha cases and 39.3% of non-Alpha cases as symptomatic (21). One of the studies in England (16) included data on “symptoms present” in a Table, without analyzing it, reporting 85.5% of Alpha cases with symptoms, and 86.6% for non-Alpha cases.

**Table 2:** Summary of symptomatic ratio data for Alpha VOC compared to wildtype.

| Location | Participants | Symptomatic ratio (95% CI) |  | Risk of bias (Newcastle-Ottawa Scale) |  |  |
| --- | --- | --- | --- | --- | --- | --- |
|  |  | Wildtype | Alpha (or proxy) | Selection | Comparability | Exposure |
| Europe (23) | 23 343 | 0.814 | 0.726 | ◇◇◇ |  | ◇◇◇ |
| Southern Italy (21) | 3075 | 0.393 | 0.517 | ◇◇◇ |  | ◇◇◇ |
| England 2 (16) | 839 278 | 0.814 | 0.855 | ◇◇◇ |  | ◇ |
| England 3 (17) | 1 767 914 | No effect across multiple symptoms |  | ◇◇ | ◇◇ | ◇ |
| England 4 (18) | 3583 | 0.4167 (0.4005, 0.4330) | 0.5292 (0.5127, 0.5456) | ◇◇◇ |  | ◇◇ |

### 3.2 Gamma

Only one study specifically analyzing Gamma was screened in, finding an overall adjusted odds ratio of 2.2 (1.8, 2.9) or 4.2 (2.1, 8.4), for Gamma, depending on the choice of technique (23). Newcastle-Ottawa Scale rating for this outcome is the same as in Table 1. The study reports a statistically significant modification of adjusted odds ratio by age, giving for CAF age groups of interest, suggesting a particularly dramatic impact amongst younger ages, although with notably wide confidence intervals:

- 20-39: 13.1 (6.5, 26.5)
- 40-59: 3.0 (1.5, 5.8)

As noted above, a study using Ontario data estimated an overall odds ratio of 1.74 (1.62, 1.86) for hospitalization for individuals with N501Y+ variants (Alpha, Beta, Gamma) relative to non-VOC (20). As Gamma at any given time during the study period made up well less than 10% of N501Y+ cases in Ontario (25), this is at best weak evidence of the specific impact of Gamma.

No studies of symptomatic ratio for Gamma were screened in. While (23) records a symptomatic ratio for Gamma, it notes that cases with available information on this variable were too rare to allow for comparisons, so it is excluded for purposes of this outcome.

### 3.3 Additional analysis

A cross-sectional study conducted in Northwest Spain involving 116 131 participants was screened in (22), although it did not report relative effects, and compared between time periods with different VOC prevalence rather than directly between cases with known VOC. We re-analyzed the data therein to look for any apparent age effect on hospitalization in the periods before (March-December 2020) and after (January-March 2021) the introduction of Alpha. In all age bands the percentage of individuals who were hospitalized decreased in the later time period, indicating that to the extent Alpha may have increased the likelihood of hospitalization, it was more than counteracted by improvements in treatment protocols or some other unknown factor. There was no clear trend in the change between age groups, at least among adults, although the drop in hospitalization rate among those under 20 was noticeably larger in percentage terms. As this study did not separate out cases between Alpha and wildtype, it is not possible to draw further conclusions; indeed, the only information on the prevalence of Alpha given in the paper is in the abstract, where it is stated that it represented 90% of new cases in Galicia in “April 2021,” while the data presented in the paper covers only up to 1 April 2021. The authors assessed this study as receiving six stars (2/1/3) on the Newcastle-Ottawa Scale.

## 4. Discussion

The primary limitation of this review was the paucity of studies of the Gamma VOC. The inconsistency of reported symptomatic ratios for VOCs and the wildtype, where they were included, makes it difficult to draw any firm conclusions; at least some of this is due to the many different methods for collecting reports of symptoms and which symptoms are or are not included.

The firmest conclusion that can be drawn from this review is that the Alpha variant comes with a higher relative risk of hospitalization compared to the original wildtype, most likely above 1.4, while unlikely to be above 2, with the balance of evidence being that the relative risk is not significantly modified by age – suggesting that general estimates of relative risk for this variant are acceptable. The evidence for Gamma is more limited, but the odds ratio may be above 2, and potentially much greater than that, especially for those 20-39 years of age (23).

In the Canadian context, the increased transmissibility of the Delta variant compared to both Alpha and Gamma (4), has it rapidly becoming the dominant variant country-wide (26). Therefore, further research on Alpha and Gamma may be less relevant to policy-makers, except to the extent that future research on the virulence of Delta may compare it to Alpha or other variants with which it is co-circulating, rather than to the original wildtype. That said, one of the studies cited here compares the hospitalization rate of Delta to the original wildtype (20); its authors estimate an odds ratio of 2.05 (1.80, 2.33) for hospitalization from Delta, compared to 1.74 (1.62, 1.86) for a mix of Alpha and Gamma (dominated by Alpha).

## Supporting information

PRISMA 2009 Checklist

ICMJE COI-AS

ICMJE COI-MRM

## Data Availability

All data used is in the cited papers.

## Authors Statement

AS: Methodology, formal analysis, investigation, resources, data curation, writing – original draft, writing – review and editing. MRM: Conceptualization, validation, formal analysis, resources, writing – original draft, writing – review and editing, supervision, project administration.

## Acknowledgements

The authors acknowledge the input and feedback of Dr. Steve Schofield on early drafts of this work, and in identifying an appropriate publication venue. Thanks are also due to Cdr Vincent Beswick-Escanlar for his feedback and support.

## References

(1) World Health Organization. Tracking SARS-CoV-2 variants [Internet]. Geneva: World Health Organization; c2021 [cited 30 Jul 2021]. Available from: https://www.who.int/en/activities/tracking-SARS-CoV-2-variants/

(2) Horby P, Bell I, Breuer J, Cevik M, Challen R, Davies N, et al. Update note on B.1.1.7 severity, 11 February 2021 [Internet]. London: New and Emerging Respiratory Virus Threats Advisor Group; 2021 [Cited 2021 Jul 30]. Available from: https://assets.publishing.service.gov.uk/government/uploads/system/uploads/attachment_data/file/982640/Feb_NERVTAG_update_note_on_B.1.1.7_severity.pdf

(3) Tuite AR, Fisman DN, Odutayo A, Bobos, P, Allen, V, Bogoch, II, et al. COVID-19 Hospitalizations, ICU Admissions and Deaths Associated with the New Variants of Concern [Internet]. Toronto: Ontario Science Table; 2021 [Cited 20201 Jul 30]. Available from: https://covid19-sciencetable.ca/sciencebrief/covid-19-hospitalizations-icu-admissions-and-deaths-associated-with-the-new-variants-of-concern/

(4) Finlay C, Archer B, Laurenson-Schafer H, Jinnai Y, Konings F, Batra N, et al. Increased transmissibility and global spread of SARS-CoV-2 variants of concern as at June 2021 [Rapid communicaiton]. Euro Surveill. 2021 Jun 17;26(24). pii: 2100509 doi: 10.2807/1560-7917.ES.2021.26.24.2100509

(5) Cevik M, Mishra S. SARS-CoV-2 variants and considerations of inferring causality on disease severity [comment]. Lancet Infect Dis. 2021. Jun 22;S1473-3099(21)00338-8. doi: 10.1016/S1473-3099(21)00338-8.

(6) Moher D, Liberati A, Tetzlaff J, Altman DG, for the PRISMA Group. Preferred reporting items for systematic reviews and meta-analyses: The PRISMA statement. BMJ. 2009;339:332–336.

(7) MacLeod MR, Hunter DG. The impact of age demographics on interpreting and applying population-wide infection fatality rates for COVID-19, INFORMS Journal of Applied Analytics, 2021;51:167–178.

(8) Biron K, Drouin PL, Guillouzic S., MacLeod MR, Schofield S, Jonasson, J. Analytic input to relaxation of pre-embarkation protocols for Naval vessels.Ottawa: Defence Research and Development Canada; 2021. Scientific Letter: DRDC-RDDC-2021-L147.

(9) Guillouzic S, Mirshak R, Sirjoosingh, A. Likelihood of undetected COVID-19 infection in a group: Effect of quarantining and testing [Internet]. Ottawa: Defence Research and Development Canada; 2020 [Cited 2021 Jul 30] Available from: http://diaenterprisepublic.canadacentral.cloudapp.azure.com/COVID19MissedInfections.html

(10) Cochrane Community, Tools and software: Covidence [Internet], London: The Cochrane Collaboration; 2021 [Cited 2021 Jul 30] Available from: https://community.cochrane.org/help/tools-and-software/covidence

(11) Fraser N, Brierley L, Dey G, Polka JK, Pálfy M, Nanni, F, et al. The evolving role of preprints in the dissemination of COVID-19 research and their impact on the science communication landscape. PLoS biology. 2021;19(4):e3000959. doi: 10.1371/journal.pbio.3000959

(12) Elsevier BV, Welcome to Scopus Preview [Internet]. Amsterdam: Elsevier; 2021 [Cited 2021 Jul 30]. Available from: https://www.scopus.com/home.uri

(13) National Center for Biotechnology Information, PubMed.gov [Internet]. Bethesda, MD: National Library of Medicine; 2021 [Cited 2021 Jul 30] Available from: https://pubmed.ncbi.nlm.nih.gov/

(14) Wells GA., Shea B, O’Connell D, Peterson J, Welch V, Losos M, et al. The Newcastle-Ottawa Scale (NOS) for assessing the quality of nonrandomised studies in meta-analyses [Internet], Ottawa: Ottawa Hospital Research Institute; 2021 [Cited 30 Jul 2021]. Available from: http://www.ohri.ca/programs/clinical_epidemiology/oxford.asp

(15) Patone M, Thomas K, Hatch R, San Tan P, Coupland C, Liao W, et al. Analysis of severe outcomes associated with the SARS-CoV-2 Variant of Concern 202012/01 in England using ICNARC Case Mix Programme and QResearch databases. medRxiv 2021.03.11.21253364 [Preprint]. 2021 [cited 2021 Jul 30]. Available from: https://www.medrxiv.org/content/10.1101/2021.03.11.21253364v1 doi: 10.1101/2021.03.11.21253364

(16) Nyberg T, Twohig KA, Harris RJ, Seaman SR, Flannagan J, Allen H et al. Risk of hospital admission for patients with SARS-CoV-2 variant B.1.1.7: cohort analysis, BMJ. 2021 Jun 15; 373:n1412. doi:10.1136/bmj.n1412

(17) Graham MS, Sudre CH, May A, Antonelli M, Murray B, Varsavsky T, et al. Changes in symptomatology, reinfection, and transmissibility associated with the SARS-CoV-2 variant B. 1.1. 7: an ecological study. The Lancet Public Health. 2021 May 1;6(5):e335–45.

(18) Walker S, Pouwels KP, House T, Coronavirus (COVID-19) Infection Survey: characteristics of people testing positive for COVID-19 in England, 27 January 2021 [Internet]. London: Office of National Statistics; 2021 [Cited 2021 Jul 30] Available from: https://www.ons.gov.uk/peoplepopulationandcommunity/healthandsocialcare/conditionsanddiseases/articles/coronaviruscovid19infectionsinthecommunityinengland/characteristicsofpeopletestingpositiveforcovid19inengland27january2021

(19) Bager P, Wohlfahrt J, Fonager J, Rasmussen M, Albertsen M, Michaelsen TY, et al. Risk of hospitalisation associated with infection with SARS-CoV-2 lineage B.1.1.7 in Denmark: an observational cohort study. Lancet Infect Dis. Forthcoming 2021, doi: 10.1016/S1473-3099(21)00290-5

(20) Fisman D, Tuite AR. Progressive increase in virulence of novel SARS-CoV-2 variants in Ontario, Canada from December to July, 2021. medRxiv 2021.07.05.21260050 [Preprint]. 2021 [cited 2021 Jul 30]. Available from: https://www.medrxiv.org/content/10.1101/2021.07.05.21260050v1 doi: 10.1101/2021.07.05.21260050

(21) Loconsole D, Centrone F, Morcavallo C, Campanella S, Sallustio A, Accogli M, et al. Rapid spread of the SARS-CoV-2 Variant of Concern 202012/01 in Southern Italy (December 2020-March 2021), Int J Environ Res Public Health. 2021 Apr 29;18(9):4766. doi: 10.3390/ijerph18094766

(22) Area I, Lorenzo H, Marcos PJ, Nieto, JJ. One Year of the COVID-19 Pandemic in Galicia: A Global View of Age-Group Statistics during Three Waves. Int. J. Environ. Res. Public Health 2021; 18(10):5104. doi: 10.3390/ijerph18105104

(23) Funk T, Pharris A, Spiteri G, Bundle N, Melidou A, Carr M,et al, Characteristics of SARS-CoV-2 variants of concern B.1.1.7, B.1.351 or P.1: data from seven EU/EEA countries, weeks 38/2020 to 10/2021, Euro Surveill. 2021 Apr 22;26(16). pii: 2100348. doi:10.2807/1560-7917.ES.2021.26.16.2100348

(24) Brown KA, Gubbay J, Hopkins J, Patel S, Buchan SA, Daneman N, et al. S-Gene Target Failure as a Marker of Variant B. 1.1.7 Among SARS-CoV-2 Isolates in the Greater Toronto Area, December 2020 to March 2021 [Research letter]. JAMA. 2021 May 25;325:2115–6.

(25) Public Health Ontario. Estimating the Prevalence and Growth of SARS-CoV-2 Variants in Ontario using Mutation Profiles [Internet]. Toronto: Queen’s Printer for Ontario; 2021 [Cited 2021 Jul 30]. Available from: https://www.publichealthontario.ca/-/media/documents/ncov/epi/covid-19-prevalence-growth-voc-mutation-epi-summary.pdf?sc_lang=en

(26) Public Health Agency of Canada. COVID-19 daily epidemiology update [Internet], Ottawa: Government of Canada; 2021 [Cited 30 Jul 2021]. Available from: https://health-infobase.canada.ca/covid-19/epidemiological-summary-covid-19-cases.html

